# An Adjudication Protocol for Severe Bacterial and Viral Pneumonia

**DOI:** 10.1101/2022.10.26.22281461

**Authors:** Chiagozie I. Pickens, Catherine A. Gao, James M. Walter, Jacqueline M. Kruser, Helen K. Donnelly, Alvaro Donayre, Katie Clepp, Nicole Borkowski, Richard G. Wunderink, Benjamin D. Singer, The NU SCRIPT Study Investigators

## Abstract

**Rationale:** Clinical endpoints that constitute successful treatment in severe pneumonia are difficult to ascertain and vulnerable to bias. Utility of a protocolized adjudication procedure to determine meaningful endpoints in severe pneumonia is not well described.

**Objectives:** To develop and validate a protocol for classification and adjudication of clinical endpoints in severe bacterial and viral pneumonia in a prospective cohort of critically ill, mechanically ventilated patients.

**Methods:** Each episode of pneumonia was independently reviewed by two of six pulmonary and critical care physician adjudicators. If a discrepancy in at least one critical answer occurred between the two adjudicators, a third adjudicator reviewed the case and answered the specific question(s) for which there was a lack of agreement. If discrepancy remained after all three adjudications, consensus was achieved through committee review.

**Results:** Evaluation of 784 pneumonia episodes during 593 hospitalizations achieved a 79% crude rate of interobserver agreement defined as agreement between 2 of 3 reviewers. Culture-negative pneumonia was associated with increased interobserver agreement. Multiple episodes of pneumonia and bacterial and viral co-infection in the initial episode of pneumonia were associated with decreased interobserver agreement. For bacterial pneumonia, patients with an adjudicated day 7-8 clinical impression of cure for the initial episode of pneumonia were more likely to be discharged alive compared to patients with a day 7-8 clinical impression of indeterminate (p < 0.01), superinfection (p = 0.03), or a combined impression of persistence and superinfection (p = 0.04). In viral pneumonia, patients with an adjudicated clinical impression of cure for an initial episode of viral pneumonia were more likely to be discharged alive compared to patients with an adjudicated clinical impression of persistence (p < 0.01), indeterminate (p < 0.01), or bacterial superinfection (p < 0.01).

**Conclusions:** We developed and validated a protocol for classification and adjudication of clinical endpoints in severe pneumonia. This protocol can be applied to cohorts of patients with severe pneumonia to provide uniform assessment of patient-centered endpoints.

## Introduction

In severe pneumonia, endpoints such as death or extubation failure are easily ascertained, while other endpoints like clinical cure, microbiologic resolution, improvement in oxygenation and safety are not standardized and subject to bias in interpretation.^1^ In studies with complex endpoints of interest, particularly events that require some analysis of clinical information, study protocols use adjudication committees to provide consistent, unbiased determination of the presence or absence of a diagnosis or an event.^2–4^ Adjudication committees are established components of large cohort studies and randomized clinical trials in cardiovascular disease, oncology, interstitial lung disease and sepsis.^2,5–9^

The process of adjudicating clinical endpoints in a large cohort of patients with severe pneumonia is not well described, although it is frequently incorporated into critical care trials.^10^ Existing studies that leverage an adjudication committee for pneumonia do not describe the process or the reliability of the method in detail.^11–13^ For example, recurrent pneumonia was utilized as an endpoint in a study of microbiological treatment failure in bacterial pneumonia, but the adjudication process used to ascertain the endpoint was not outlined.^14^ A large randomized controlled trial (RCT) of community-acquired pneumonia (CAP) employed an endpoint adjudication committee to determine the primary outcome, clinically indicated treatment with antibiotics, but did not detail the validity of the adjudication process.^15^ Other literature focuses on diagnostic adjudication in pneumonia and does not address endpoint adjudication. For example, a study by Cook et al reported a crude agreement rate ranging from 50% to 82% when two adjudicators evaluated ventilator-associated pneumonia (VAP) in patients enrolled in a randomized controlled trial; however, this study was performed almost 25 years ago and focused on VAP, limiting the generalizability of these findings to a broader patient population with severe pneumonia.^16^ A 2013 study by Klouwenberg et al described interobserver agreement between clinicians identifying the source of infection and the plausibility of infection in critically ill patients with sepsis using the Centers for Disease Control and Prevention criteria, finding the lowest rate of interobserver agreement in VAP with only 35% full diagnostic agreement between two observers.^17^ The authors concluded the CDC criteria are insufficient to assess complex infections like pneumonia. Additionally, a prospective study on infection in patients with the systemic inflammatory response syndrome reported that the lowest rate of physician agreement occurred when identifying the presence of pneumonia.^18^ These studies describe the complexity of diagnosing pneumonia but do not address methods for standard identification of clinical endpoints in pneumonia.

We developed a protocol for adjudication of clinical endpoints for a prospective cohort of critically ill, mechanically ventilated patients with suspected pneumonia. We hypothesized that our procedures reliably assess endpoints and exhibit construct validity in patients with severe pneumonia. Our results suggest that our protocol is a useful and valid tool to provide uniform and consistent identification of endpoints in bacterial and viral pneumonia.

## Methods

This adjudication protocol was developed to assess episodes of pneumonia for patients enrolled in the single-center **<u>S</u>**uccessful **<u>C</u>**linical **<u>R</u>**esponse **I**n **<u>P</u>**neumonia **<u>T</u>**herapy (SCRIPT) study (Northwestern University IRB #STU00204868). SCRIPT is an NIH/NIAID-sponsored U19 Systems Biology Center that uses a multi-omics approach to analyzing pathogen, host defense and microbiome factors in order to understand the response to severe pneumonia at the alveolar level. SCRIPT is a prospective, observational cohort study of mechanically ventilated patients with suspected pneumonia. In the medical intensive care unit (ICU) at Northwestern Memorial Hospital (NMH), bronchoalveolar lavage (BAL) or non-bronchoscopic BAL (NBBAL) is routinely performed to confirm presence and identify etiology of pneumonia in intubated patients.^19–22^ In this manuscript, the term BAL will encompass both bronchoscopic and non-bronchoscopic BAL procedures. Our adjudication protocol was developed prior to the start of patient enrollment in the SCRIPT study in 2018 and subsequently modified during the COVID-19 pandemic in 2020 to include adjudication of primary viral pneumonia and viral pneumonia complicated by bacterial superinfection.

### Clinical Endpoints

Our protocol was designed to ascertain a final clinical endpoint at the time of death or hospital discharge as well as interim clinical endpoints. The interim clinical endpoints were defined as Cure, Indeterminate, Persistence or Superinfection. See the Supplement for detailed definitions of each term. For bacterial pneumonia or bacterial and viral co-infection pneumonia interim clinical endpoints were assessed at day 7-8, day 10 and day 14, corresponding to common antibiotic discontinuation time points in clinical trials and clinical practice. Endpoints were defined based on literature review and expert consensus.^23–25^ Because part of our objective was to evaluate patients at various time points during an episode of pneumonia, we did not use traditional measures like mortality, ventilator-free days or microbiological cure as stand-alone endpoints.

### Pneumonia Episode Definition

We defined the initial episode of pneumonia by the date of the clinical BAL procedure that prompted enrollment into SCRIPT. An episode of pneumonia ended when systemic antibiotics for pneumonia were discontinued without recurrent signs of pneumonia for at least 48 hours (defined in detail in the Supplement). Onset of subsequent episodes within the same hospitalization were likewise defined by the timing of a diagnostic BAL procedure occurring after at least 48 hours off antibiotics directed at pneumonia and without recurrent or worsening signs of pneumonia during the interval. We applied standard definitions for pneumonia^26,27^ at the time of each BAL procedure. See the Supplement for descriptions of clinical community-acquired pneumonia (CAP), hospital-acquired pneumonia (HAP) and ventilator-associated pneumonia (VAP) and additional definitions used to standardize the adjudication process. Importantly, the definitions of CAP, HAP and VAP were applied based on the duration of admission or ventilation preceding the time of BAL sampling irrespective of the timing or locale of symptom onset. We defined episodes on and subsequent to the date of the SCRIPT study enrollment BAL procedure up to day 99, censoring any episodes preceding SCRIPT enrollment or beginning after day 99.

### Adjudication Committee

The adjudication committee was composed of six pulmonary and critical care physicians at Northwestern Medicine. Two adjudicators independently and retrospectively reviewed each patient’s complete electronic health record (EHR) and entered written responses to each question on the evaluation form (included in the Supplement). Review was conducted after the patient’s hospital discharge. If both adjudicators provided the same responses to all questions on the evaluation form, consensus was achieved. If discrepancy in at least one response occurred, the discrepant question was highlighted on a new, blank evaluation form and given to a third adjudicator who answered the specific question based on chart review, blinded to the previous reviewers’ responses. Agreement between the third reviewer and one of the original reviewers was considered the final response. If a discrepancy remained between all three reviewers, a committee review was performed with a minimum of three adjudicators present, and a consensus answer was provided by the committee. If a case was identified by the first reviewer as a non-pneumonia episode, this classification was confirmed by a second reviewer if requested by the first reviewer. The first reviewer could request committee review for unusual, complex cases.

Committee adjudication took place in person or by video conference on a weekly basis. The total committee group membership at any time point was five physicians. The committee held several meetings for quality control to ensure the manual data entered into the adjudication worksheet aligned with the data entered into the electronic database and the designated etiologies for pneumonia (i.e., viral, bacterial, co-infection, culture-negative, non-pneumonia and indeterminate) were accurate per written guidelines available to all committee members (included in the Supplement).

### Adjudication Worksheet

The full adjudication worksheet consisted of 20 questions and is included in the Supplement. Depending on the complexity of the case and the branching logic of the form, not all questions required a response. Additionally, some questions had objective, numerical answers that did not require multi-reviewer concordance to resolve discrepancies. Because the answers to these questions could be identified by chart review, a research coordinator reviewed the case to resolve discrepant responses to these questions rather than involving a third reviewer: (1) Number of days antibiotics were given for viral-only pneumonia, (2) If initial sample, has the patient been actively treated for pneumonia for more than 24 hours prior to sample collection, (3) Are serial procalcitonin values available, and (4) Level of evidence for extrapulmonary infection.

### Statistical Analysis

Statistical analysis was performed using R Studio (RStudio 2022.07.0: Integrated Development for R. RStudio, PBC, Boston, MA). The Sankey diagram in Figure 1 was created using Python 3.9. Comorbidities were mapped using ICD codes with Charlson Comorbidity Index definitions, with only comorbidities present before admission contributing to totals. Pearson’s Chi-squared or Fisher’s exact test with Bonferroni correction for multiple comparisons was performed for outcomes with categorical variables. The conditional maximum likelihood estimator was used to report odds ratios. Crude rates of interobserver agreement were calculated. To estimate a probability of chance agreement for the Cohen’s Kappa Statistic (observed agreement - chance agreement/1-chance agreement) we calculated the probability of chance agreement on three mandatory questions: Appropriate antibiotics (3 response options), Clinical Impression (5 response options) and the Overall Global Clinical Cure (2 response options). This resulted in a 0.04 probability of chance agreement.

**Figure 1.**
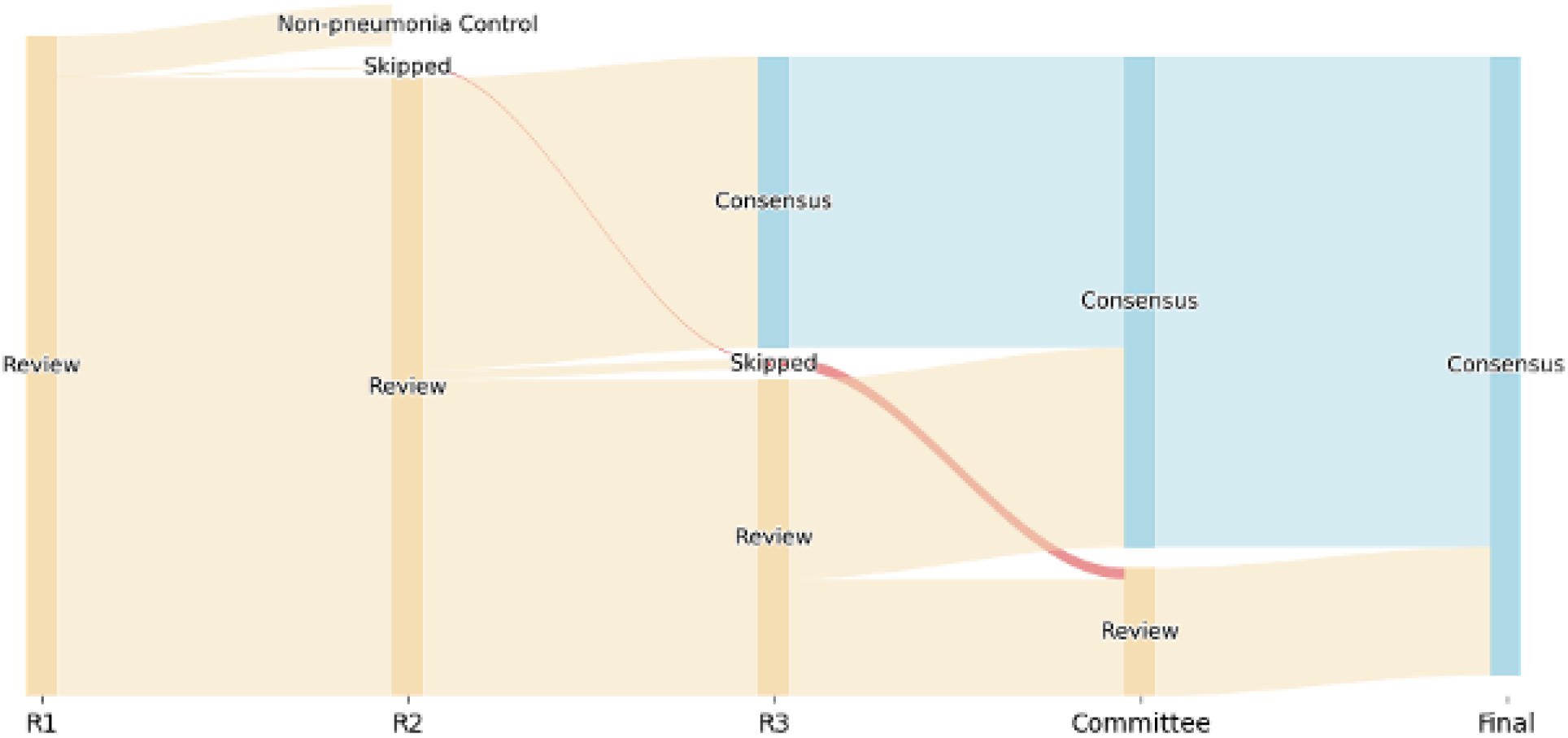
Sankey diagram illustrating the adjudication process. Out of 593 hospitalizations, 37 had only one reviewer (R1), 262 cases had two reviewers (R2), 294 cases required a third reviewer (R3). A small subset went directly to committee review after R1 and R2.

## Results

From June 2018 to June 2022, 784 pneumonia episodes from 593 hospitalizations were adjudicated. Patient demographics and clinical characteristics are listed in Table 1.

**Table 1.** Demographics of participants with adjudicated episodes of pneumonia. *Patients with viral only pneumonia, non-pneumonia and those who were deceased, discharged or transferred to another facility prior to day 7-8 did not have a day 7-8 adjudication.

| **Variables** | **N (%)** |
| --- | --- |
| Age (Median [IQR]) | 62 [51,72] |
| **Gender** |  |
| Male | 298 (59.1%) |
| Female | 206 (40.9%) |
| **Ethnicity** |  |
| Hispanic | 104 (20.6%) |
| Not Hispanic or Latino | 377 (74.8%) |
| Not answered | 23 (4.6%) |
| **Race (Self-Identified)** |  |
| African-American or Black | 100 (19.8%) |
| American Indian or Alaska Native | 4 (0.8%) |
| Asian | 17 (3.4%) |
| Native Hawaiian or Other Pacific Islander | 1 (0.2%) |
| White | 297 (58.9%) |
| Not answered | 85 (16.8%) |
| **Pneumonia Category for Initial Episode (n = 593)** |  |
| Clinical CAP | 137 (23.1%) |
| Clinical HAP | 202 (34.1%) |
| Clinical VAP | 160 (27.0%) |
| Non-pneumonia episode (37 single reviewer, 57 adjudicated) | 94 (15.9%) |
| **Pneumonia Etiology (n = 519)** |  |
| Viral only | 153 (25.8%) |
| Bacterial only | 150 (25.3%) |
| Viral/bacterial co-infection | 87 (14.7%) |
| Indeterminate | 3 (0.1%) |
| Culture-negative | 106 (17.9%) |
| **Number of Episodes (n = 593)** |  |
| Multiple episodes | 122 (20.6%) |
| Single episode | 471 (79.4%) |
| **Comorbid Conditions** |  |
| Cerebrovascular disease | 124 (20.9%) |
| Congestive heart failure | 173 (29.2%) |
| Malignancy | 200 (33.7%) |
| Diabetes | 213 (35.9%) |
| Chronic pulmonary disease | 212 (35.7%) |
| Liver disease | 155 (25.2%) |
| Renal disease | 159 (26.8%) |
| **Hospitalization** |  |
| Hospital length of stay (median, interquartile range [IQR]) | 23 [13,37] |
| ICU length of stay (median, [IQR]) | 14 [6,25] |
| Patients with day 7-8 adjudication* | 257 (60.0%) |
| Hospital mortality | 189 (37.5%) |

37/593 (6.2%) hospitalizations were not reviewed by multiple adjudicators because the first reviewer identified the episode(s) as non-pneumonia or requested an expedited committee review for unusual or previously undefined conditions; thus the total number of hospitalizations reviewed by multiple adjudicators was 556 (Figure 1). 262/556 (47.1%) hospitalization-level examinations had an interobserver concordance between two adjudicators and did not require a third adjudicator; hence, 294/556 (52.9%) patient cases went to a third reviewer to evaluate at least one point of disagreement. 117 of the 239 cases that required a third reviewer had a discrepancy between all three adjudicators and required a committee review to arrive at a final agreement on the appropriate endpoint(s) for the patient. Thus, overall consensus (either agreement of 2/2 or 2/3 reviewers) without the need for committee review was achieved in 439/556 (78.9%) cases.

To identify differences in the rates of consensus agreement over time, we calculated crude rates of interobserver agreement by year (Figure 2). At least two reviewers adjudicated 41 cases from 2018, 110 cases from 2019, 258 cases from 2020, 124 cases from 2021 and 23 cases from 2022. In the years spanning 2018 to 2022, interobserver agreement defined as agreement between two of two adjudicators was 54%, 51%, 41%, 52% and 57% respectively. The Cohen’s Kappa coefficient per year for interobserver agreement based on agreement between two of two adjudicators was 0.52, 0.49, 0.39, 0.50 and 0.55, respectively. When interobserver agreement was defined as agreement between 2 of 3 adjudicators, the crude rate of agreement increased to 85.4%, 85.5%, 69.8%, 86.3% and 100% agreement. A test for proportions demonstrated a significant difference in interobserver agreement by year (X^2^ = 27.1, df = 4, p = < 0.001). A test for proportions excluding 2020 demonstrated no significant difference in interobserver agreement between 2018, 2019, 2021 and 2022 (X^2^ = 3.8, df = 3, p = 0.28), reflecting the temporal reliability of the adjudication protocol outside of the early pandemic year 2020.

**Figure 2.**
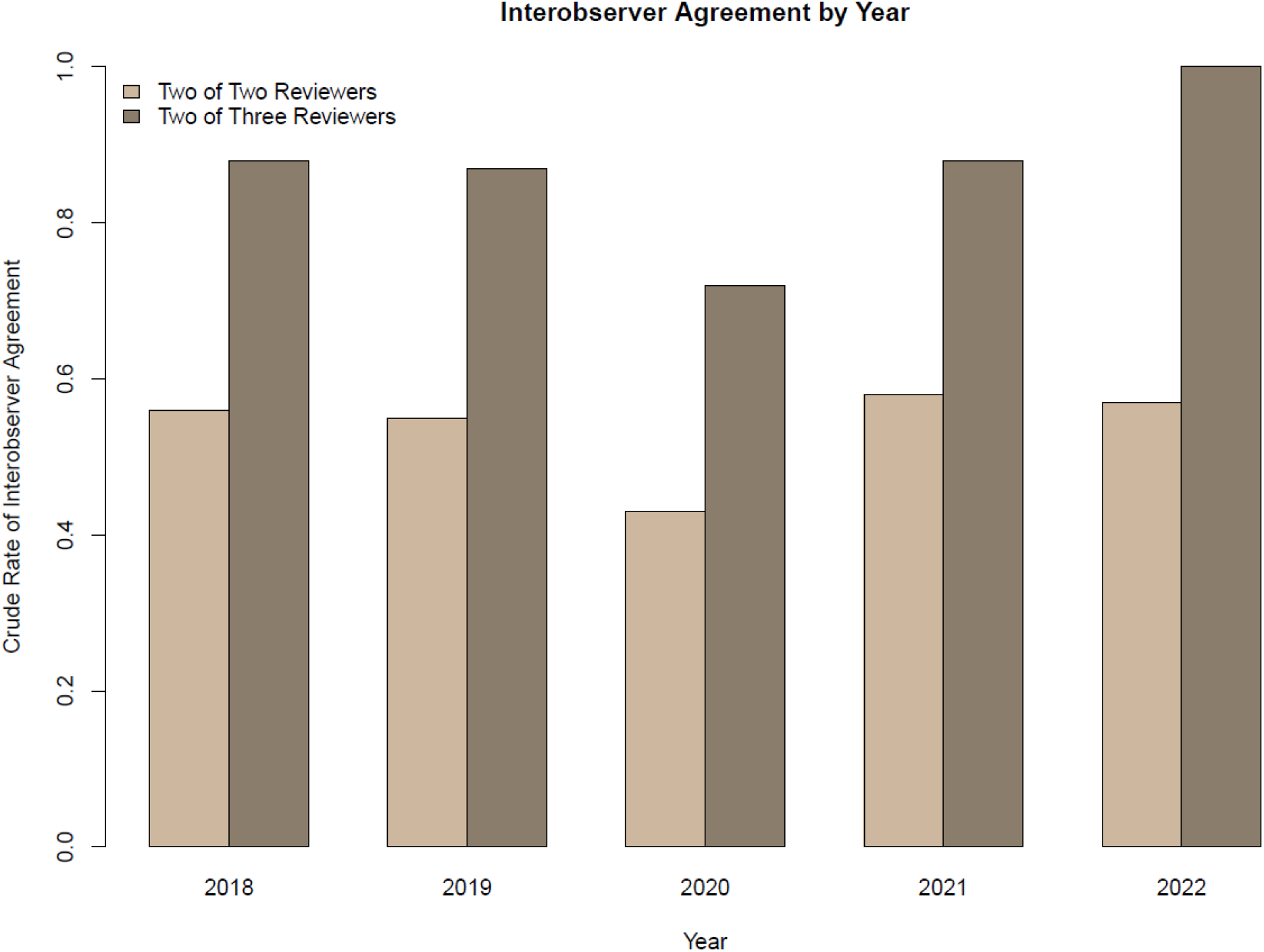
Crude rates of interobserver agreement by year. When interobserver agreement was defined as consensus between 2 of 2 reviewers, the crude rates of interobserver agreement did not exceed 0.6. Interobserver agreement defined as agreement between 2 of 3 reviewers increased the rate of agreement.

We sought to determine whether the etiology of the initial pneumonia episode and the number of pneumonia episodes in a given hospitalization were associated with interobserver agreement, defined as consensus between 2 of 3 reviewers without the need for committee review. The proportion of initial pneumonia episodes that were CAP, HAP or VAP did not differ between cases that had consensus without committee review and cases that required committee review for consensus. The odds ratio for consensus from 2 of 3 reviewers for CAP was 0.78 (95% CI [0.49, 1.24]), for HAP was 1.17 (95% CI [0.75, 1.85]) and for VAP was 0.76 (95% CI [0.48 to 1.21]).

We calculated the odds of interobserver agreement based on the microbiologic etiology of pneumonia. For example, for pneumonia with a defined bacterial etiology, we divided the odds of interobserver agreement among the cases with an initial episode of pneumonia with a defined bacterial etiology by the odds of interobserver agreement among the cases where the initial episode was not bacterial pneumonia. An odds ratio > 1 suggests a higher proportion of interobserver agreement in the adjudication worksheet and less need for committee review of at least one of the critical elements of the adjudication worksheet. Initial episodes of pneumonia that were bacterial and viral co-infection were associated with lower interobserver agreement (OR = 0.57, 95% CI [0.33, 0.98]) (Figure 3). Initial episodes of pneumonia that were culture-negative were significantly associated with higher interobserver agreement (OR = 2.15, 95% CI [1.13, 4.36]). The presence of multiple episodes was strongly associated with a committee review required to achieve consensus (OR = 0.13, 95% CI [0.08, 0.20]). Initial episodes of pneumonia that were viral only did not have a significant difference in the odds of interobserver agreement versus committee review to achieve consensus (OR = 0.64, 95% CI [0.40, 1.01]).

**Figure 3.**
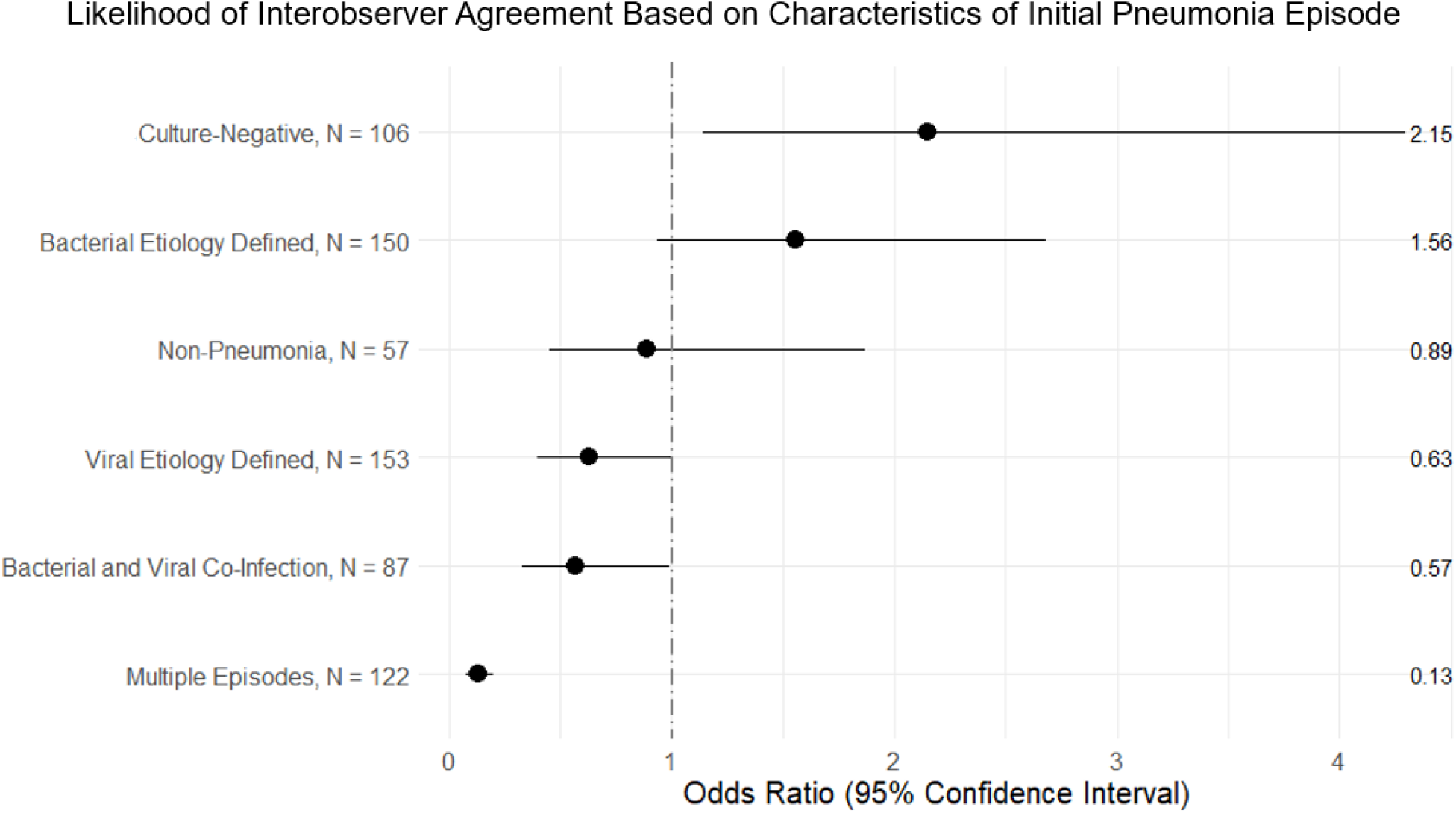
The odds of interobserver agreement based on characteristics of the initial episode of pneumonia.

To determine the construct validity of our clinical endpoints, we assessed the discharge disposition based on the day 7-8 adjudicated clinical endpoint of the initial pneumonia episode. A day 7-8 adjudicated clinical endpoint was available for 311 hospitalizations; 282 did not have a day 7-8 clinical endpoint for the initial episode because 94 were non-pneumonia episodes, 153 had viral infection only and 35 were discharged, transferred to another hospital or died prior to a day 7-8 assessment. Discharge dispositions were grouped into favorable outcomes (home, acute inpatient rehabilitation, skilled nursing facility, long term acute care facility) versus unfavorable outcomes (death or hospice). As demonstrated in Figure 4, the proportion of patients discharged to favorable versus unfavorable dispositions differed by the day 7-8 clinical endpoint of the initial episode of pneumonia.

**Figure 4.**
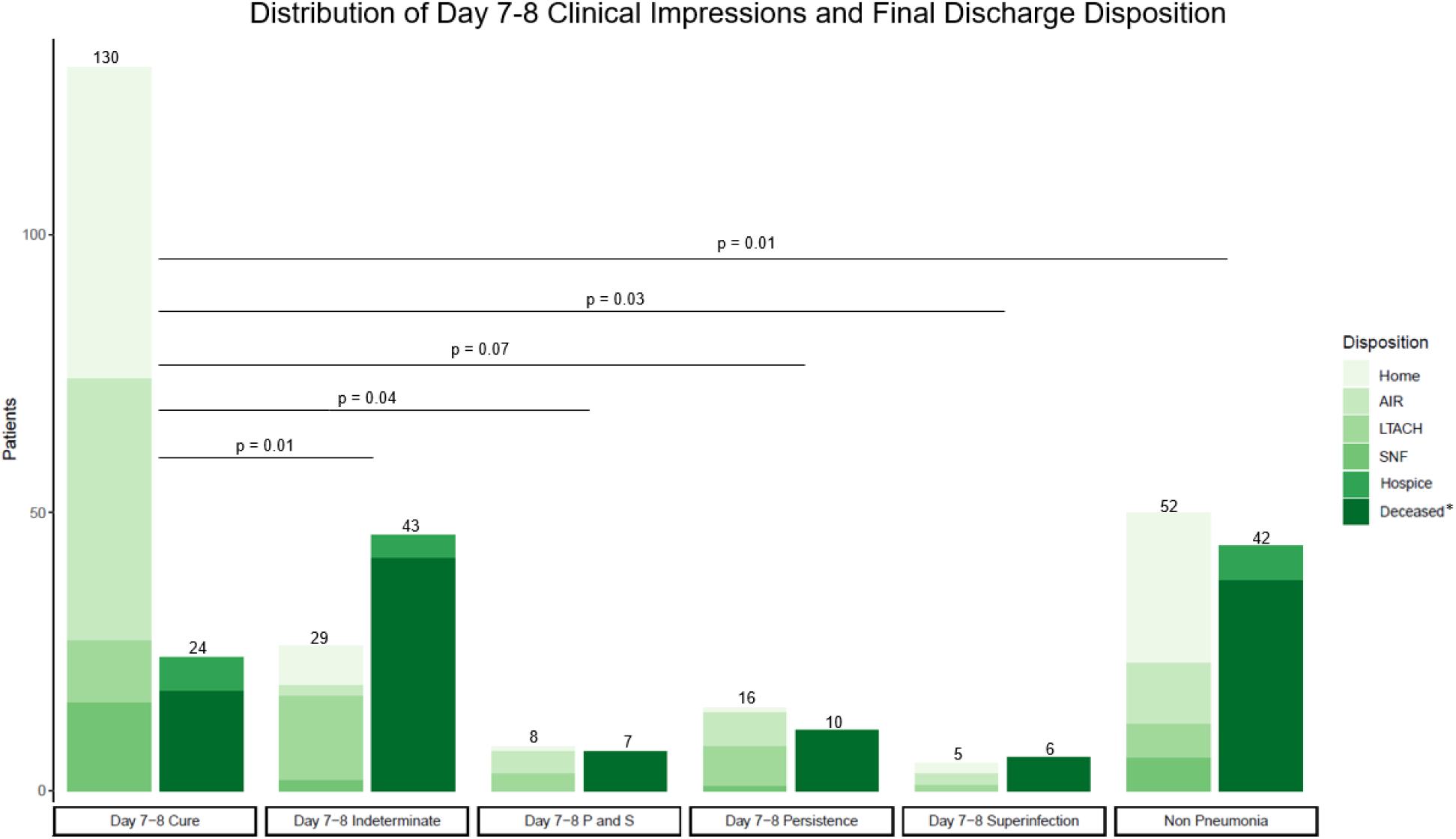
The association between day 7-8 clinical endpoints (cure, indeterminate, persistent or superinfection) for the initial episode of bacterial or bacterial-viral co-infection pneumonia and discharge disposition at the end of the hospitalization. Day 7 P and S is an abbreviation for Persistence and Superinfection. *Deceased includes patients who underwent lung transplantation for refractory respiratory failure during their hospitalization. Lines with p values compare mortality rates between groups.

A higher proportion of patients with a day 7-8 clinical endpoint of cure had a favorable discharge disposition compared to patients with a day 7-8 clinical endpoint of indeterminate, superinfection or persistence with superinfection. No significant difference in discharge disposition was found when cure was compared to persistence alone. As demonstrated in Table 2, in patients with viral only pneumonia a higher proportion of patients with a clinical impression of cure were discharged to favorable dispositions compared to patients with an adjudicated clinical endpoint of indeterminate, superinfection or persistence.

**Table 2.** Differences in hospital discharge disposition based on the adjudicated clinical endpoints of patients with severe viral pneumonia.

| Clinical Endpoints for Initial Episode of Viral Pneumonia | Control Group (Cure) Discharge Disposition |  | Comparison Group Discharge Disposition |  | p Value |
| --- | --- | --- | --- | --- | --- |
|  | Home, AIR, SNF, LTACH | Deceased or Hospice | Home, AIR, SNF, LTACH | Deceased or Hospice |  |
| Cure | 47 | 3 | -- | -- | NA |
| Persistence | -- | -- | 0 | 19 | <0.01 |
| Indeterminate | -- | -- | 0 | 15 | <0.01 |
| Superinfection | -- | -- | 37 | 30 | <0.01 |

## Discussion

We developed and validated an adjudication protocol for severe bacterial and viral pneumonia in intubated patients. Pneumonia is a complex clinical syndrome with poorly defined endpoints for treatment success or failure. Because of this, the FDA suggests all-cause mortality at 14 to 28 days as a primary endpoint for noninferiority registration clinical trials of new therapy for bacterial HAP and VAP.^28^ However, mortality fails to capture important patient-centered outcomes.^29^ Using the Delphi method, a panel of international experts attempted to establish a consensus composite endpoint for clinical trials related to HAP and VAP.^20^ The panel discussed a range of possible endpoints, including clinical cure, ventilator-free days, decrease in clinical pulmonary infection score (CPIS), microbiological cure, safety, change in procalcitonin and acquisition of antimicrobial resistance. The range of proposed endpoints highlight the complexity that clinicians and researchers face when evaluating whether a patient with pneumonia had a favorable or unfavorable outcome. In the aforementioned study by Weiss et al, clinical cure was ranked as the most important primary outcome, yet a universally accepted definition for clinical cure in pneumonia does not exist. Weiss and colleagues used an iterative process to ultimately define clinical cure as resolution of signs and symptoms of infection, improvement in oxygenation and no appearance of new signs of sepsis. In this study, we incorporate these clinical characteristics into our definition of cure at day 7-8, day 10 and day 14.

Our protocol for clinical classification and adjudication of episodes of pneumonia resulted in an 79% overall crude rate of consensus agreement on clinical endpoints when two adjudicators independently reviewed episodes of pneumonia and a third adjudication was available to resolve discrepancies. Based on literature review, our consensus agreement rate falls within the realm of “strong” or “substantial” agreement.^30^ Sjoding et al reported a similar finding in which the addition of a third reviewer improved interobserver reliability for adjudication of complex clinical cases of ARDS.^31^ The extent of our adjudication worksheet minimizes the probability that two observers could provide the same responses to all of the questions by chance. In a highly cited review, McHugh et al advise researchers to rely on the kappa statistic if there is likely to be much guessing between observers, but if raters are well trained and there is low likelihood of guessing, it is safe to rely on percent agreement as a reflection of interrater reliability.^32^

In our study, the presence of multiple episodes of pneumonia during a hospitalization was associated with lower odds of consensus by interobserver agreement. An initial episode of bacterial/viral co-infection pneumonia also decreased the odds of consensus by interobserver agreement. This finding likely reflects the increased complexity of assessing co-infection, particularly when microbiologic resolution for one pathogen and persistence of the other exists. Conversely, interobserver agreement was significantly higher for culture-negative cases. The reason for this is unclear but may be related to the fact that there are few variables to consider when determining whether or not the infection has resolved in culture negative cases. For example, decrease in colony forming units of an organism does not apply to culture-negative cases.

We observed a statistical decrease in agreement for episodes occurring in 2020 during the early portion COVID-19 pandemic, in which our adjudication methods for viral pneumonia were changed to accommodate the high numbers of SARS-CoV-2 pneumonia cases in our cohort. Use of a new instrument that required iterative clarification of definitions in the midst of a large increase in cases enrolled contributed to the higher rates of discrepancies. In addition, patients with SARS-CoV-2 pneumonia had complex clinical courses, which may have challenged objective identification of disease resolution.

For patients with bacterial pneumonia, we identified an association between day 7-8 adjudicated clinical endpoint of cure and discharge from the hospital alive compared to a day 7-8 adjudicated clinical endpoint of indeterminate, superinfection or combined persistence and superinfection. For patients with viral pneumonia, we identified an association between a clinical endpoint of cure and discharge from the hospital alive compared with an adjudicated clinical endpoint of indeterminate, superinfection or persistence. This important association between adjudicated endpoints and objective outcomes supports the construct validity of our procedures and suggests that hospital mortality is linked with the clinical course of severe pneumonia at day 7-8 of the pneumonia episode.

Our study has limitations. We did not obtain input from independent observers, such as the physicians treating the patient, to benchmark the results from the adjudication committee, although we did review all clinical documentation from the bedside teams during our case reviews. Clinical adjudication committees are controversial and some studies report no difference in the rate of events reported by individual observers compared with the committee. Another limitation is the external validity of our findings, which may depend on the experience of the adjudicators comprising the panel. A lower agreement rate may result from clinicians less familiar with the diagnosis and management of severe pneumonia. In our study, key data incorporated into adjudications came from BAL fluid results, which may limit the generalizability of our protocol in centers where BAL sampling is used less often in pneumonia diagnosis. Nevertheless, our classification and adjudication protocol serve as a useful starting point for investigators to define clinical endpoints in severe bacterial and viral pneumonia.

## Supporting information

NU SCRIPT Investigators

## Data Availability

All data produced in the present study are available upon reasonable request to the authors.

## Acknowledgements

The authors would like to acknowledge Nikolay Markov for programming assistance.

## Notes

**Funding:** National Institutes of Health, National Institute of Allergy and Infectious Diseases (AI135964), Northwestern Memorial Hospital Masters in Medicine Institutional Grant. CAG is supported by NIH awards T32HL076139 and F32HL162377. JMK is supported by NIH/NHLBI K23HL146890. RGW is supported by NIH grants U19AI135964, U01TR003528, P01HL154998, R01HL14988, R01LM013337. BDS is supported by NIH awards R01HL149883, R01HL153122, P01HL154998, P01AG049665, and U19AI135964.

**Declaration of interests:** BDS holds US patent 10,905,706, “Compositions and methods to accelerate resolution of acute lung inflammation,” and serves on the Scientific Advisory Board of Zoe Biosciences, for which he holds stock options. Other authors declare no conflicting interests.

### Competing Interest Statement

BDS holds US patent 10,905,706, Compositions and methods to accelerate resolution of acute lung inflammation, and serves on the Scientific Advisory Board of Zoe Biosciences, for which he holds stock options. Other authors declare no conflicting interests.

### Funding Statement

National Institutes of Health, National Institute of Allergy and Infectious Diseases (AI135964), Northwestern Memorial Hospital Masters in Medicine Institutional Grant. CAG is supported by NIH awards T32HL076139 and F32HL162377. JMK is supported by NIH/NHLBI K23HL146890. RGW is supported by NIH grants U19AI135964, U01TR003528, P01HL154998, R01HL14988, R01LM013337. BDS is supported by NIH awards R01HL149883, R01HL153122, P01HL154998, P01AG049665, and U19AI135964.

### Author Declarations

Northwestern University Institutional Review Board gave ethical approval for this work (#STU00204868).

### Summary of Updates

The middle initial has been added to author's name. Figure 4 has been added to the manuscript.

