## Supplement for "An Adjudication Protocol for Severe Bacterial and Viral Pneumonia"

| **Definitions** | |
|---|---|
| Pneumonia Episode | In our institution, BAL sampling is performed for clinical suspicion of pneumonia. Lower respiratory tract infection (pneumonia) is diagnosed by positive BAL culture, positive BAL multiplex PCR or negative BAL culture with a white blood cell differential containing greater than 50% neutrophils and no alternative explanation. Episodes begin on the day of the BAL procedure and end when antibiotics for pneumonia are discontinued for at least 48 hours and clinical signs and symptoms of pneumonia are improving. |
| Pneumonia Episode Category Assessment | **Viral only:** A virus is detected from an upper or lower respiratory tract specimen with evidence of inflammation in BAL fluid.<br><br>**Bacterial/viral co-infection:** A virus is detected from an upper or lower respiratory tract specimen and bacteria by positive BAL fluid culture or PCR.<br><br>**Bacterial only:** A bacteria is detected from BAL by positive culture or positive PCR with negative viral detections from upper and lower respiratory tract samples or not tested.<br><br>**Culture-negative:** No viral or bacterial organisms are detected from respiratory specimens but the BAL fluid white blood cell differential contains greater than 50% neutrophils and no alternative explanation is likely.<br><br>**Indeterminate:** The BAL culture is negative and no cell count or differential is available on the BAL fluid specimen in a patient with clinical signs and symptoms of pneumonia. |
| Pneumonia Episode Category | **Clinical CAP**: Pneumonia diagnosed at the time of BAL, when BAL was performed within 48 hours of hospitalization if the patient was not hospitalized within the last 7 days.<br><br>**Clinical HAP:** Pneumonia diagnosed at the time of BAL, when BAL was performed after 48 hours of hospitalization or if the patient was discharged from a healthcare facility within the last 7 days where admission was greater than 24 hours.<br><br>**Clinical VAP:** Pneumonia diagnosed at the time of BAL, when BAL was performed after at least 48 hours of mechanical ventilation or if the patient was reintubated less than 24 hours after extubation.<br><br>**Non-pneumonia episode:** Clinical data and respiratory sample results not consistent with pneumonia (negative BAL culture and BAL fluid differential contains less than 50% neutrophils). Examples include aspiration pneumonitis, pulmonary hemorrhage, heart failure or non-pneumonia-induced ARDS. |
| Appropriate Antibiotics | The organism(s) identified by culture or multiplex PCR was susceptible to the initially prescribed antibiotics. |

| | |
|---|---|
| Clinical Endpoints for Viral Pneumonia | The outcome of an episode of viral only pneumonia.<br><br>**Cure:** Documented clearance of the virus by PCR or clinical resolution of signs of infection and acute respiratory failure. Extubation without subsequent reintubation within 48 hours represents cure.<br><br>**Persistence:** The viral PCR remains positive within 7 days of death or transfer out of the hospital with persistent lung or systemic inflammation.<br><br>**Superinfection pneumonia:** The patient has an initial episode of viral pneumonia and a new episode of secondary bacterial pneumonia.<br><br>**Indeterminate:** If nasopharyngeal swab (NP) is positive but BAL is negative, no repeat/recent PCR within 7 days prior to death or transfer to another facility. |
| Clinical Endpoints for Bacterial Pneumonia or Bacterial and Viral Co-Infection Pneumonia | The status of a patient at day 7-8, 10 and 14. Day 10 and day 14 were not adjudicated if antibiotics were stopped at an earlier time point.<br><br>**Cure:** (all of the following must be present to consider cure) 1) The patient survived the duration of antibiotic treatment specifically for pneumonia, 2) antibiotics for pneumonia were stopped without recurrent/superinfection pneumonia within 48 hours of discontinuation or the same antibiotics were continued for more than 14 days but signs/symptoms of pneumonia were stable (WBC, secretions, oxygenation, radiograph) or improving, 3) the causative pathogen disappeared from respiratory secretions or no further samples were obtained, 4) the clinical manifestations of pneumonia improved/resolved (e.g., fever, secretions, WBC, hypoxemia, septic shock) and 5) the patient was able to wean from the vent, initiate spontaneous breathing trials or return to pre-pneumonia ventilator/ECMO settings. Extubation without subsequent reintubation within 48 hours represents cure.<br><br>**Persistence:** Positive BAL culture or PCR, blood, pleural fluid, or endotracheal aspirate culture of same pathogen; abscess/cavity; empyema; or endocarditis.<br><br>**Superinfection bacterial or viral pneumonia:** New pneumonia with new bacterial/viral pathogen during treatment for the episode and the patient was not off antibiotics for more than 48 hours.<br><br>**Indeterminate**: BAL neutrophilia, persistent fever, elevated WBC without defined etiology, persistent indication of inflammation but |

| | |
|---|---|
| | no respiratory correlates for failure of pneumonia treatment (hypoxemia, secretions, worsening radiograph). |
| Final Clinical Endpoint | The final clinical outcome was assessed at the time of hospital discharge or death.<br><br>**Success:** Survival for the duration of treatment or beyond with at least one of the following: (1) ability to stop antibiotics without recurrence or superinfection pneumonia (2) causative pathogen disappears from respiratory secretions or no further samples obtained (3) clinical manifestations of pneumonia improve or resolve (4) ability to wean from the ventilator or initiate spontaneous breathing trials.<br><br>**Failure:** Death while still being treated for pneumonia, complications from pneumonia (i.e. empyema or abscess) or persistent hemodynamic instability until a change in antibiotics. The potential causes of failure are listed below, in Section 8 titled "Definitions For Cause of Failure." |

**PNEUMONIA EPISODE OUTCOME ADJUDICATION GUIDE (SECTIONS 1-8)**

Episode start date is linked to first SCRIPT BAL, not the start of fever/leukocytosis/abnormal imaging, or start of antibiotics, etc.

**Section 1.**
**Actively Treated Prior to Initial Enrollment Sample**

- Y or N within 24 hours **prior** to BAL sample.
- If Y, count by calendar days, (first day of ABX is 0).
  - Ex: piperacillin-tazobactam started 7/1 at 22:20, BAL on 7/4, number of antibiotic days = 3
- If known, include PNA treatment from outside hospital for relevant episode.
- If OSH records not available, refer to NMH records only (check pharmacy notes as well)
- If an ongoing antibiotic regimen was changed to target pneumonia prior to initial BAL, count days starting on date in which ABX were changed to cover pneumonia
  - Ex: initially treated for non-pulmonary infection, new fever -> suspected HAP -> ABX changed -> initial study BAL taken. Start counting the day pneumonia-targeted ABX were started
  - If patient is treated for sepsis with unknown source, only start counting date in which ABX for pneumonia coverage were started
- For viral pneumonia patients, count by date in which ABX for suspected bacterial pneumonia coverage were started. These would not be considered prophylaxis.

**Section 2.**
**Appropriate Initial Antibiotics**

- Antibiotics in the 72 hours preceding culture susceptibility results.
- No antibiotics until culture results known would be considered inappropriate
- May use BioFire or other Dx testing results with sensitivity or resistance genes known, if applicable.
- If uncommon pathogen or BioFire positive only, assume usual antibiotic susceptibility

**If Multiple Episodes:**

- Must have a **$\geq$ 48-hour period off systemic antibiotics for previous episode or no systemic antibiotics for pneumonia** prior to start of a new episode

**Section 3.**
**Viral only Episode**

- Were appropriate antivirals administered? At any time during this viral infection episode, even if started as outpatient/outside hospital.
- Should receive most of the usual course e.g., >= 4 days remdesivir.
- Any antiviral with FDA indication or EUA is considered appropriate.
- Retreatment with an alternative agent at the time of a new episode of pneumonia should be reported, e.g., baloxivir in a patient with prior treatment with oseltamivir
- N/A should be checked if no antivirals clinically active against the virus exist.

**Section 4.**
**Bacterial/Viral Co-Infection**

- If prior viral only episode, viral testing information carries over. No need to repeat information and N/A may be selected for second and subsequent episodes.
- If persistence of viral is retreated with same or alternative agent, record for “appropriate anti-viral”
- Add new virus if identified at subsequent episode, “Were appropriate antivirals administered?” should be answered only for new virus.

**Section 5.**
**Viral Pneumonia Clinical Endpoint/Final Viral Clinical Endpoint**

**Complete Global Viral Clinical Endpoint if Viral only episode (first four are mutually exclusive – i.e., check only one):**
Pertains to virus information only throughout the study.

- Cure- cleared virus (documented by PCR) or clinical resolution of signs of infection and acute respiratory failure
- Persistence, if viral PCR positive within 7 days of death or transfer to LTAC with persistent inflammation
- Superinfection pneumonia, only if more than 1 episode and new bacterial pneumonia
- Indeterminate, if NP positive but BAL clear, no repeat/recent (within 7 days) PCR prior to death or transfer to other facility
- Extrapulmonary infection if single viral episode only-check appropriate bacterial not pneumonia site

**Section 6.**
**Bacterial (including Culture-negative) Pneumonia Clinical Endpoint Criteria (Days 7, 10, 14 if applicable)**
**Cure: yes to all the criteria below**

1. survive duration of treatment (or beyond)
2. able to stop antibiotics without recurrent/superinfection pneumonia within 48 hours of discontinuation
    a. Continuation of same antibiotics >14 days **<u>but</u>** signs/symptoms of pneumonia stable (WBC, secretions, oxygenation, radiograph) or improving
3. causative pathogen disappears from respiratory secretions or no further samples
    a. %PMNs in repeat BAL < 50% in non-neutropenic
4. clinical manifestations of pneumonia improve/resolve (e.g., fever, secretions, WBC, hypoxemia, septic shock)
5. ability to wean from vent or at least initiate SBTs
    a. Drop in minute ventilation and/or improvement in oxygenation
    b. Patient returned to pre-pneumonia ventilator/ECMO status and/or weaned from vent

**Consider failure if any of the below:**

- Did the patient die while still being treated with antibiotics for pneumonia?
- Did development of pneumonia directly lead to family/POA decision to shift to comfort care?
- Complications (empyema/abscess)
- Did the patient have persistent need for vasopressors or hemodynamic instability until a change in antibiotics?

**Persistence**

- Positive BAL culture or PCR, Blood, pleural fluid, or ETA culture of same pathogen
- Abscess/Cavity, Empyema, Endocarditis, or Other (not extrapulmonary superinfection)

**Superinfection bacterial or viral pneumonia**: new pneumonia with new bacterial/viral pathogen during treatment for this episode.

- Superinfection and original pathogen results carried forward for final episode determination
- Not off antibiotics for > 48 hours during treatment
- If > 48 hours off antibiotics, consider a new episode
- Not considering fungal/Mycobacterial/parasitic superinfections

**Indeterminate (for clinical endpoint only)**: BAL neutrophilia, persistent fever, and leukocytosis without defined etiology

- Persistent indications of inflammation but no respiratory correlates for failure of pneumonia treatment (hypoxemia, secretions, worsening radiograph)
- BAL % PMNs > 50%
- Extrapulmonary infection with possible pneumonia coverage by antibiotic

**Section 7.**
**Overall Global Clinical Cure Criteria**

The clinical picture at the end of the study. The end of the study is defined as death, discharge, or lung transplant.

**Success: all criteria below**

1. survive duration of treatment (or beyond)
2. able to stop antibiotics for pneumonia
   a. Continuation of same antibiotics > 14 days but signs/symptoms of pneumonia stable (WBC, secretions, oxygenation, radiograph) or improving?
3. causative pathogen disappears from respiratory secretions or no further samples
   a. %PMNs in repeat BAL < 50% in non-neutropenic
4. clinical manifestations of pneumonia improve/resolve (e.g., fever, secretions, WBC, hypoxemia, septic shock)
5. ability to wean from vent or at least initiate SBTs
   a. Drop in minute ventilation and/or improvement in oxygenation
   b. Patient returned to pre-pneumonia ventilator/ECMO status or weaning from vent

**Success without all criteria:**

Pneumonia was treated and considered cured but did not meet all criteria outlined

**Consider "Failure" if any of the below:**

- Did the patient die while still being treated for pneumonia?
- Did development of pneumonia directly lead to family/POA decision to shift to comfort care?
- Complications (empyema/abscess/endocarditis) uncontrolled at time of end of study
- Did the patient have persistent need for vasopressors or hemodynamic instability until end of study?
- Would patient have likely died of persistent pneumonia without intervention of lung transplant? Should then be on antibiotics specific to pneumonia until time of transplant (may have additional/change for other infections if still covering pneumonia pathogen). If antibiotics for pneumonia are stopped, still cure if antibiotics needed for other site of infection. If no other

site identified i.e.. “sepsis” but antibiotic coverage for VAP pathogens = failure for persistent inflammation

**Section 8.**
**Definitions for cause of Failure**

**Antibiotics for other indication**

- Receiving antibiotics initially started for pneumonia, but continued for extrapulmonary infection, that could cover pneumonia at time of study exit

**Persistence bacterial pneumonia**

- Continued BAL, blood, or ETA positive culture or BAL PCR of same pathogen within 7 days of end of study
- Ongoing antibiotic treatment for Abscess/Cavity, Empyema, Endocarditis, or Other at end of study

**Persistent inflammation only**

- BAL neutrophilia, persistent fever, WBC w/o defined etiology
- Death beyond duration of treatment but from sepsis or MOSF that started with onset of pneumonia
- Persistent indications of inflammation but no respiratory correlates (hypoxemia, secretions, worsening radiograph)
- BAL % PMNs > 50%
- If viral only episode, PCR positive within 7 days of end of study and viral pneumonia should be limiting ability to wean from mechanical ventilation or persistent inflammation and respiratory failure without alternative infectious diagnosis

**Recurrence**

- New pneumonia episode with same bacterial pathogen after treatment cure for prior episode but treatment for recurrence not completed prior study exit
- Positive culture/PCR and restart of antibiotics >48 hours of stop
- Signs and symptoms of pneumonia clearly resolved in the interval.

**Superinfection pneumonia**

- New pneumonia with **new** pathogen that arises during treatment for another pathogen.
- High % PMNs without positive culture or BioFire should NOT be considered superinfection. Designate as persistent inflammation.
- Only used if being treated at the end of enrollment. Otherwise, would new episode or continuation or previous episode.
- Not considering fungal/Mycobacterial/parasitic superinfections. If these are suspected cause of death, code as persistent inflammation
